# Presence and vitality of SARS-CoV-2 virus in wastewaters and rivers

**DOI:** 10.1101/2020.05.01.20086009

**Authors:** Sara Giordana Rimoldi, Fabrizio Stefani, Anna Gigantiello, Stefano Polesello, Francesco Comandatore, Davide Mileto, Mafalda Maresca, Concetta Longobardi, Alessandro Mancon, Francesca Romeri, Cristina Pagani, Lorenzo Moja, Maria Rita Gismondo, Franco Salerno

## Abstract

Wastewater-based epidemiology has been proposed to monitor the diffusion and trend of SARS-CoV-2 pandemic. In the present study, raw and treated samples from three wastewater treatment plants, and two river samples characterized the Milano Metropolitan Area, Italy, were surveyed for SARS-CoV-2 RNA positivity to real time PCR and infectiveness. Moreover, whole genome sequencing and phylogenetic analysis of isolated strains was performed.

Raw wastewater samples resulted positive to PCR amplification, while treated water samples were always negative (four and two samples, respectively, sampled in two dates). Moreover, the rate of positivity in raw wastewater samples decreased after eight days, in congruence with the epidemiological trend estimated for the interested provinces. Virus infectiveness was always not significant, indicating the effectiveness of wastewater treatments, or the natural decay of viral vitality, which implied the absence of significant risk of infection from wastewaters. Samples from receiving rivers (two sites, sampled in the same dates as wastewaters) showed in some cases a positivity to PCR amplification, probably due to non-treated discharges, or the combined sewage overflows. Nevertheless, also for rivers vitality was negligible, indicating the absence of sanitary risks. Phylogenetic analysis of genome indicated that the isolated virus belongs to the most spread strain present in Europe and similar to another strain found in Lombardy.

## 1. Introduction

On March 11, 2020, the World Health Organization (WHO) has officially declared the novel coronavirus (COVID-19) outbreak a global pandemic. This emergency saw an unprecedented investment of energies and resources by the scientific community to rapidly contains the spread of the virus. Surveillance of SARS-CoV-2 circulation and real time trend monitoring were advocated as key actions to activate pandemic responses. The high rate of asymptomatic infected individuals (Al-Tawfiq, 2020) has challenged the estimation of infection spread basing on survey on ill patients, and alternative approaches, such as wastewater-based epidemiology, were proposed (Medema et al., 2020; Randazzo et al., 2020). Since SARS-CoV-2 duration in faecal samples of infected patients is supposed to be high (Gupta et al., 2020; Zhang et al., 2020), the identification and quantification of viral RNA in raw wastewater (WW) could be a reliable marker of infection prevalence in the population indeed, as demonstrated for other viruses (Hovi et al., 2012; Wigginton et al., 2015). Coronaviruses, however, are enveloped viruses, and their persistence in the environment could be short, although little is known concerning SARS-CoV-2 survival in water (Naddeo and Liu, 2020). Moreover, little is known also about the potential diffusion of this virus in the aquatic environment, mediated by the WW collection and treatment network. Similarly, vitality test of SARS-CoV-2 in treated WW has shown that in some cases virus could be potentially still infectious (Wurtzer et al., 2020).

In Europe, northern Italy was one earliest and most infected area to date, with about 200’000 cases on 27 April 2020 (https://www.ecdc.europa.eu/en/geographical-distribution-2019-ncov-cases). The northern part of Italy has reported the prevalence of Italian cases. In particular the Milano Metropolitan Area, including the Province of Milano and Monza/Brianza with 18’559 and 4’516 cases, respectively, experienced an infection rate (case divided by resident population) almost double (0.58 and 0.52%, respectively) respect to the rest of Italy (0.32%) *(https://github.com/pcm-dpc/COVID-19/blob/master/dati-province/dpc-covid19-ita-province.csv)*. In the present study, we evaluated the presence of SARS-CoV-2 RNA in raw and treated WWs collected in three wastewater treatment plants (WWTPs), covering the entire Milano Metropolitan Area, including the northeastern densely inhabited productive area and the Milano urban centre. The main aims of this survey were: 1) evaluating the effectiveness of WWTPs in reducing or eliminating the viral load, after checking for the presence of SARS-CoV viral RNA in WWs; 2) testing for virus vitality before and after WWTPs, in relation to the intrinsic persistence of SARS-CoV-2; 3) comparing genomic strains of SARS_CoV2 found in WWs to those isolated from ill patients in the area and worldwide; 4) checking for the presence of SARS-CoV viral RNA in rivers downstream the Milano Metropolitan Area.

The results here discussed are part of a more extended activity, aimed at evaluating the use of wastewater-based epidemiology for the complex case of the Milano Metropolitan Area. WWs are currently being sampled and preserved during the decreasing phase of the epidemy, and more sensitive techniques will be tested and fine tuned in the meanwhile, basing on this first survey.

## 2. Materials and Methods

### 2.1 Study area, sample collection and filtration

Samples have been collected in three WWTPs, one of them located in the Province of Monza/Brianza (named WWTP-A) and two of them located in the Province of Milano (named WWTP-B and WWTP-C). All these three WWTPs globally collect 11 m^3^/s of sewage from approximately 2 million inhabitants (Figure 1). These facilities are all equipped with secondary treatments and a tertiary disinfection step by peracetic acid or high intensity UV lamps. The WWTP-A and WWTP-B discharge into the Lambro River, while the WWTP-C into the Lambro Meridionale River (Figure 1). Both rivers and all WWTPs were sampled in the same two dates: April, 14^th^ and April, 22^th^, 2020 almost at the same hour (1.00 p.m., - instantaneous samples-). Samples were stored in polypropylene (PP) 500 mL bottles and immediately transferred under refrigeration to laboratory for filtration. Grab samples of rivers were collected from river bridges using a stainless steel bucket and transported in dark glass bottles to laboratory under refrigeration. Water samples were pre-filtered on glass fiber filters (Whatman GF/F, 0.7 μm nominal pore size, 145 mm diameter), then on Millipore 0.2μm nominal pore size, 145 mm diameter filters. All glassware was disinfected by an ipochlorite solution between every sample process. Being the assay of vitality one of the main aim of this work, we prefer to avoid any preliminary concentration treatment of samples, without assessing the effects on virus vitality, since it is likely that the addition of chemical compounds or mechanical stress could hamper the survivorship and infectiveness of coronaviruses (Gundy et al., 2009; Ye et al., 2016).

**Figure 1.**
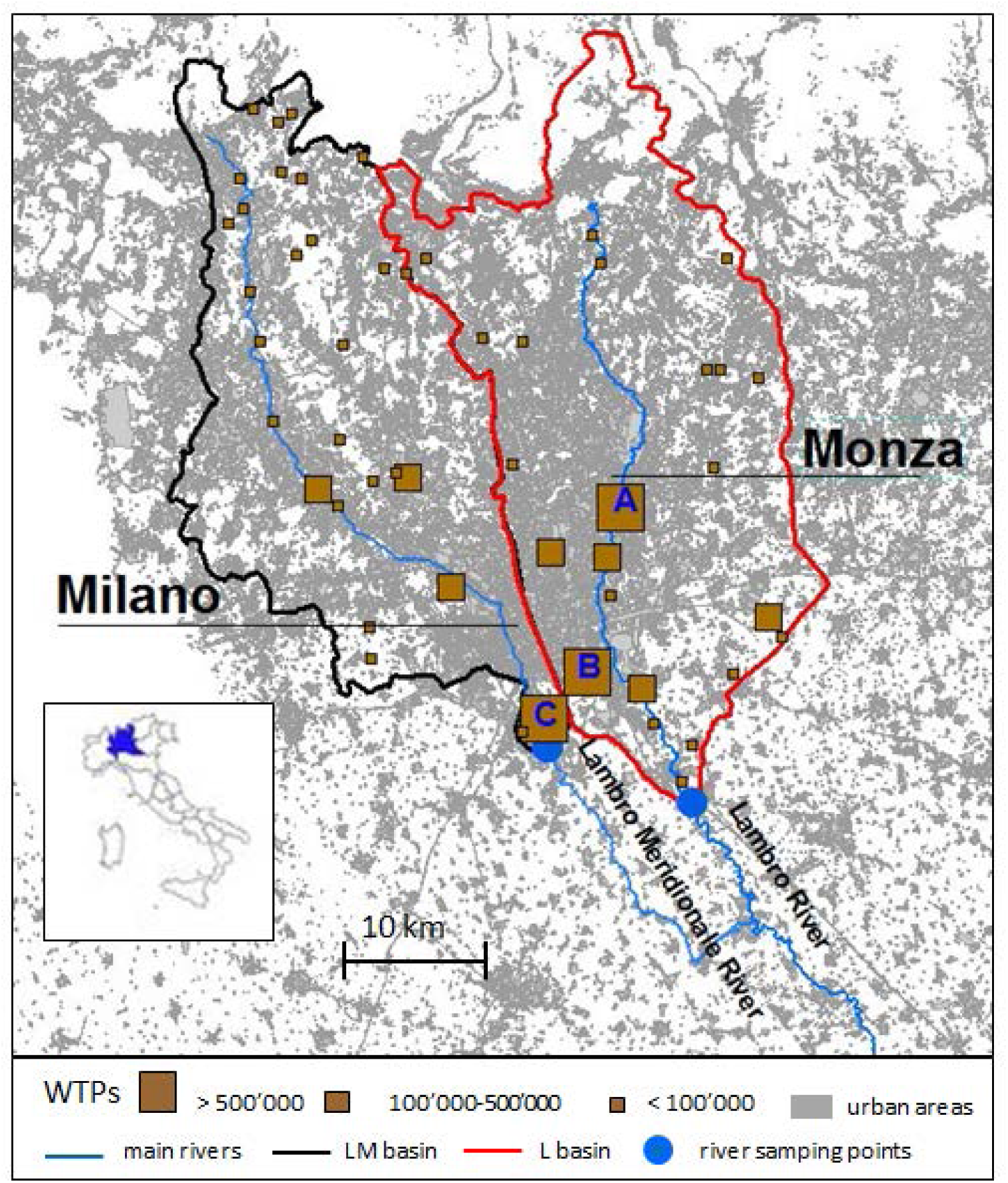
Map of Milano Metropolitan Area, with the two main river basins highlighted: 1) the Lambro Meridionale (LM) River, closed and sampled on Mirasole bridge; 2) the Lambro (L) River, closed and sampled on Melegnano bridge. Locations of all WWTPs are indicated with squares whose size is proportional to their capacity (population equivalent). WWTP-A is the main plant of Monza/Brianza. WWTP-B and WWTP-C are the main plants of Milano. These WWTPs have been investigated in this study.

### 2.2 SARS-CoV-2 RNA extraction and real time PCR

Detection protocols already available for diagnostic routine were employed in the present case, to answer the demand for the rapid survey of WWs samples during the epidemic peak. A total of 200 μl of filtered samples was used for viral RNA extraction using the QIAMP VIRAL RNA mini kit (Qiagen, Hilden, Germany), using standard protocols indicated by the manufacturer.

To detect and assess the presence of the SARS-CoV-2, 5 μl of RNA was tested with a qualitative commercial real-time PCR (Liferiver, Shanghai, China). SARS-CoV-2 presence was assessed by using the 2019-nCoV Real-Time RT-PCR Diagnostic Panel contains primers and probes that target the nucleocapsid (N) gene (designed for both universal detection of SARS-like coronaviruses as well as specific detection of six 2019-nCoV strains), the ORF1ab gene and the E gene.

### 2.3 Whole genome sequencing

Twenty microliters of the obtained genomic RNAs were retro-transcribed using 2 random primers pools (Promega, Italy, dettagli). 10 μl of c DNA was sequenced by Ion Torrent (Thermofisher, Monza, Italy) according to the manufacturer’s instructions. The Ion Ampliseq library kit plus was used for library preparations and whole genome sequencing was performed accordingly.

### 2.4 Genome assembly and phylogenomic analysis

Genome assembly was obtained using a mapping-based approach. Low quality read bases were trimmed out using Trimmomatic software with the MAXINFO:50:0.3 parameter set. Then, SNP calling was performed using the Wuhan-Hu-1 strain genome (accession MN908947.3) as reference. The genome consensus sequence was obtained on the basis of the identified SNPs and the reference sequence. Reference bases were called in conserved positions with coverage above five, otherwise N were introduced.

A dataset of 3,995 SARS-CoV-2 genomic sequences were retrieved from GISAID database (Elbe and Buckland-Merrett, 2017). The multi-genome alignment including our genome, GISAID genomes and reference genome was obtained using the Purple pipeline (Gona et al., 2020), (available at https://skynet.unimi.it/index.php/tools/purple-tool/). The nucleotide distances among the strains were computed using the R library Ape and the 300 most similar sequences to our strain were selected for Maximum Likelihood phylogenetic analysis. The phylogenetic analysis was performed using RaxML8 software with 100 pseudo bootstraps, after model selection using ModelTest-NG. The obtained phylogenetic tree was visualized using iTol web tool.

### 2.5 Cell culture and virus isolation

In order to evaluate the vitality of SARS-CoV-2, viral isolation protocol was conducted through the utilisation of cell culture, which included VERO E6 cells (ATCC® CRL-1586™), a monkey kidney cell line. VERO cells were cultured in Dulbecco’s Modified Eagle Medium with 1-glutamine (DMEM, Gibco™ ThermoFisher Scientific), these were supplemented with 10% of heat-inactivated fetal bovine serum (FBS, Gibco™ ThermoFisher Scientific) and 1% Penicillin-Streptomycin [5,000 U/mL] (Pen-Strep, Gibco™ ThermoFisher Scientific) and incubated in a 37°C incubator at 5% CO2 atmospheric pressure. The viral isolation protocol was performed for each sample. Firstly, a 25cm2 cell culture flask was used in which 2 mL of sample and 5 mL of DMEM at 2% of heat-inactivated FBS and 1% of Pen-Strep. All of which were incubated for 72h at 37 °C with CO2 level of 5% were inoculated. At the end of the 72h wait, 3 mL of cells culture supernatant was inoculated into a new 25cm2 cell culture flask, together with 5 mL of medium (DMEM at 2% FBS and 1% Pen-Strep) and again incubated at 37 °C with a CO2 level of 5% for 48h. Finally, vitality was assessed daily by screening cells for for cytopathic effects.

## 3. Results and discussion

### 3.1 Presence and vitality of SARS-CoV-2 in wastewaters

The amplification of SARS-CoV-2 RNA genes ORF1ab, N and E was successful in the raw WWs from all the WWTPs on April, 14^th^, 2020, and only in the raw WW of the WWTP-B plant on April, 22^nd^, 2020. On the contrary, treated WWs did not result positive to real time PCR on any date (Table 1). Results suggest the effectiveness of the treatments to reduce the viral load up to some order of magnitude, being all treated samples negative to RNA amplification. However, we cannot exclude that viral copies were still present in the outflow of the WWTPs at low concentrations, under the sensitivity threshold of the multiplex reaction. Indeed, lockdown of public research activities in Italy, except for healthcare institutions, did not allow to tailor specific protocols suited to concentrate virus from environmental samples, and to perform RNA quantification, for the emerging case of COVID19.

**Table 1.** Results of real time PCR amplification, vitality test and genome sequencing, as obtained for WWTs and rivers for two dates. Genes code refers to the nucleocapsid (N) gene, the Human RNase Polymerase (Orf1ab) gene and the E gene (E). n.a.: analysis not performed

| Date | Sample origin | Station | treatment | Gene positivity |  |  | Vitality test | Genome code |
| --- | --- | --- | --- | --- | --- | --- | --- | --- |
|  |  |  |  | Orflab | N | E |  |  |
| 14/04/2020 | WWTPs | A | raw | + | - | + | No cytopathic effect | hCoV-19/Italy/HSacco-1/2020 |
|  |  | A | treated | - | - | - | No cytopathic effect | n.a. |
|  |  | B line 1 | raw | + | - | + | n.a. | n.a. |
|  |  | B line 2 | raw | + | + | + | No cytopathic effect | n.a. |
|  |  | C | raw | - | - | - | n.a. | n.a. |
|  |  | C | treated | - | - | - | n.a. | n.a. |
|  | Rivers | Lambro Meridionale | - | + | + | - | n.a. | n.a. |
|  |  | Lambro | - | + | + | - | n.a. | n.a. |
| 22/04/2020 | WWTPs | A | raw | - | - | - | No cytopathic effect | n.a. |
|  |  | A | treated | - | - | - | No cytopathic effect | n.a. |
|  |  | B line 1 | raw | + | + | - | No cytopathic effect | n.a. |
|  |  | B line 2 | raw | - | - | - | No cytopathic effect | n.a. |
|  |  | C | raw | - | - | - | No cytopathic effect | n.a. |
|  |  | C | treated | - | - | - | No cytopathic effect | n.a. |
|  | Rivers | Lambro Meridionale | - | - | - | - | No cytopathic effect | n.a. |
|  |  | Lambro | - | + | + | - | No cytopathic effect | n.a. |

Conventional WWTPs (i.e., WWTPs based on chlorine disinfection before the final release) proved to be efficient in removal viral load up to 4 log10 (Wang et al., 2018) for many viral groups. In the specific case of SARS-CoV-2 few data are available at this regard. In the WWTPs of Paris (Wurtzer et al., 2020) a 100-fold reduction of viral load after treatments was estimated as well, starting from values around 1 × 10^7^ in the raw WW. In other studies analyzing viral concentration only in the inflow to WWTPs, SARS-CoV-2 genome concentration was around 50-1500 copies/ml for the nucleocapsid genes (Nemudryi et al., 2020; Wu et al., 2020). Given a sensitivity of 1000 viral copies/ml for the multiplex PCR kit employed in this study, the observed positivity may suggest high levels of SARS-CoV-2 in the sewage of Milano Metropolitan Area, comparable to those estimated in Paris (Wurtzer et al., 2020). Nevertheless, quantitative data are not available, and the targeted genes proved different sensitivity thresholds (Jung et al., 2020), so that estimates of viral concentration should be weighted by the sensitivity of the employed marker. In this respect, ORF1ab gene showed the highest frequency of positivity, and resulted amplified in all positive samples, while both the other two genes (N and E) failed to be amplified in two out of five positive cases. We also cannot exclude that PCR inhibitors could be present in the reactions, and that targets were differently sensitive to their presence, although avoiding samples concentration should have minimized this risk.

Interestingly, the positivity disappeared in most of the inlet samples on April, 22nd, eight days apart from the first sampling, indicating a possible decrease of the viral load. Figure 2 shows the daily number of new daily cases of the two Provinces interested by this study (provided by the national survey service), normalized by the resident population in each Province. Both Provinces recorded the same magnitude of relative daily new diagnosed cases and similar trends. The infection started to increase since the end of February, 2020, reached a maximum in the second half of March, 2020, and then has slowly decreased. These epidemiological data need standardized collection methods (Sims and Kasprzyk-Hordern, 2020) and in Italy are probably affected by unequal sampling efforts and asynchrony of records respect to the real infection dates. Hence, temporal correlation between wastewater-based and clinical epidemiology can be hampered by these factors. However, in this study amplification data are congruent with the main falling trend of the epidemic event, suggesting that wastewater-based epidemiology could be a feasible approach in monitoring the diffusion and prevalence of SARS-CoV-2 infection (Mallapaty, 2020). It is worth of mention that, at this purpose, more sensitive quantitative approaches (i.e., quantitative PCR on concentrated viral samples) should be tuned.

**Figure 2.**
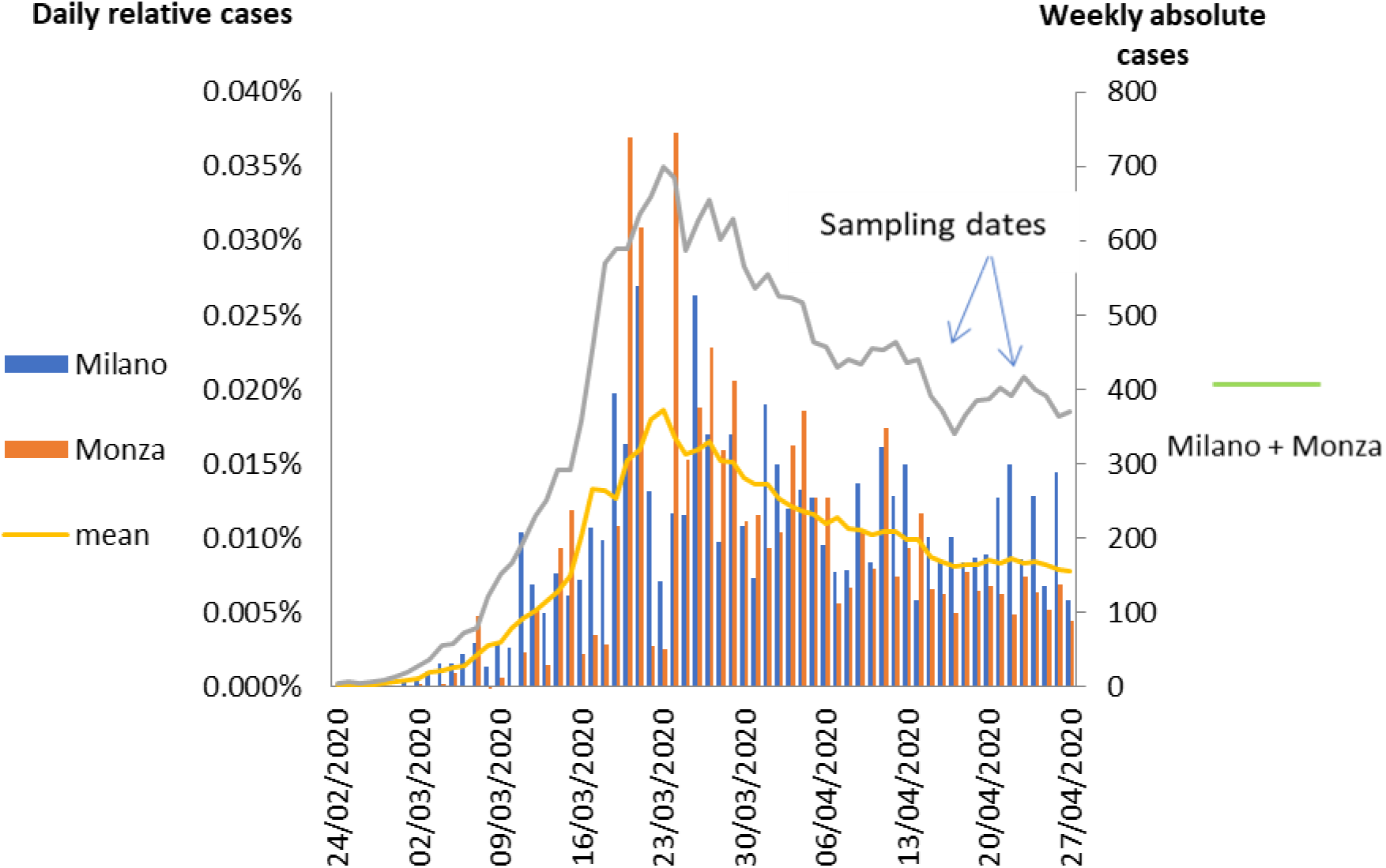
Trend of COVID-19 cases diagnosed in Province of Milano and Monza/Brianza (source: Protezione Civile, https://github.com/pcm-dpc/COVID-19/blob/master/dati-province/dpc-covid19-ita-province.csv). On the left y-axis, relative cases represent the new daily cases divided by the resident population in each Province. On the right y-axis, absolute cases represent the weekly window mean of the sum of the new daily absolute cases of both Provinces.

The vitality of SARS-CoV-2 in WWs, either raw or treated samples, resulted not significant, despite the likely high number of RNA copies present in the samples. Indeed, no cytophatic effects on Vero E6 cells were detected at 48 h and 72 h after inoculation. Enveloped virus are more susceptible to decay of their infectiveness in wastewaters than non enveloped viruses (Ye et al., 2016), especially when in presence of free active enzymes activity or predators like protozoan or metazoan (Kim and Unno, 1996). Survival of coronaviruses, or enveloped viruses, in wastewaters is also dependent on temperature, being around 7-13 h, but higher in some conditions, at mild temperature (23-25°C) and longer in colder conditions (36 h at 10°C) (Casanova et al., 2009; Ye et al., 2016). In the present case-study, the time from faeces emission to the arrival at the WWTP has been estimated to be about 6-8 hours according to the mean corrivation times of WWs provided by the WWST managers. Climatic conditions were mild (15-20°C) in the weeks of sampling activities, and probably did not favour the survival of SARS-CoV-2 up to the WWTPs. A few studies have evaluated the persistence of free viral RNA in waters, which can vary from less than one hour in WWs (Limsawat and Ohgaki, 1997) to two days in sea water (Tsai et al., 1995), depending also on environmental conditions and virus typology. Any speculation about the role of wastewater treatments in reducing the viral concentration, respect to the natural decay of virus load in the WWTPs, cannot be advanced at this stage. At this regard, specific analysis targeting the persistence of SARS-CoV-2 RNA in WWs under different environmental conditions are ongoing. Public concern for potential infection risk due to accidental contacts with WWs (e.g., airborne aerosols and droplets) seems to be negligible in the case of the investigated WWTPs. From the environmental point of view, WWTPs should not constitute a significant source of infective SARS-CoV-2.

### 3.2 Presence and vitality of SARS-CoV-2 in rivers

Positive amplification of viral RNA was found in both receiving rivers on April, 14^th^, 2020, but only in the Lambro River on April, 22^nd^, 2020 (Table 1), following probably the epidemiological decreasing trend already described for WWs. The presence of SARS-CoV-2 RNA in rivers in spite of the absence in treated WWs of the analysed plants probably indicates that an aliquot of non-treated WWs was present in the surface waters, and this situation was exacerbated in the sampling period characterised by an anomalous and prolonged drought (Bollettino Idrologico Arpa Lombardia, 2020). The explanations could be the presence of non-collected domestic discharges or the lack of separation of the urban runoff waters from the domestic effluents, which causes combined sewer overflows (CSOs). The latter possible source of contamination is common to the central Europe, with approximately 70% of combined sewer systems (Butler and Davies, 2004) and the United States, there are approximately 40 million people served by this facility (U.S. Environmental Protection Agency (EPA), 2004). Combined sewer overflows (CSOs) usually occur during high precipitation events in order to prevent damages on the WWTPs. However, even during prolonged drought, some CSO devices can be active due to possible failures of the sewerage system (Salerno et al., 2018). Nevertheless, all the possible sources should be investigated by using anthropogenic substances, such as e.g. caffeine, which are specific tracers of untreated domestic sewages (Viviano et al., 2017).

However, it is worth of mention that also in the positive case of the River Lambro the vitality of the SARS-CoV-2 was negligible, indicating the absence of sanitary and environmental risk of infection from river water.

### 3.3 Phylogenetic analysis of SARS-CoV-2 strains

A single genome was sequenced so far, associated to a strain isolated in WWTP-A. The assembly procedure allowed to call 25,279 genome bases, corresponding to ~85% of the reference genome length. Phylogenetic analysis (Figure 3) revealed that the sequenced strain is closely related to a SARS-CoV-2 strain isolated on March, 3^th^, 2020 in Milan (GISAID code EPI_ISL_413489). We found two SNPs between the two strains, including a non-synonymous mutation on the ORF1 gene at position 2231 (L2231I). Moreover, this strain is within the main clade of European genomes, congruently with a common origin. Genome sequencing of other isolates are currently ongoing, and will provide more detailed information about the occurrence of specific haplotype in the Italian population (Bai et al., 2020).

**Figure 3.**
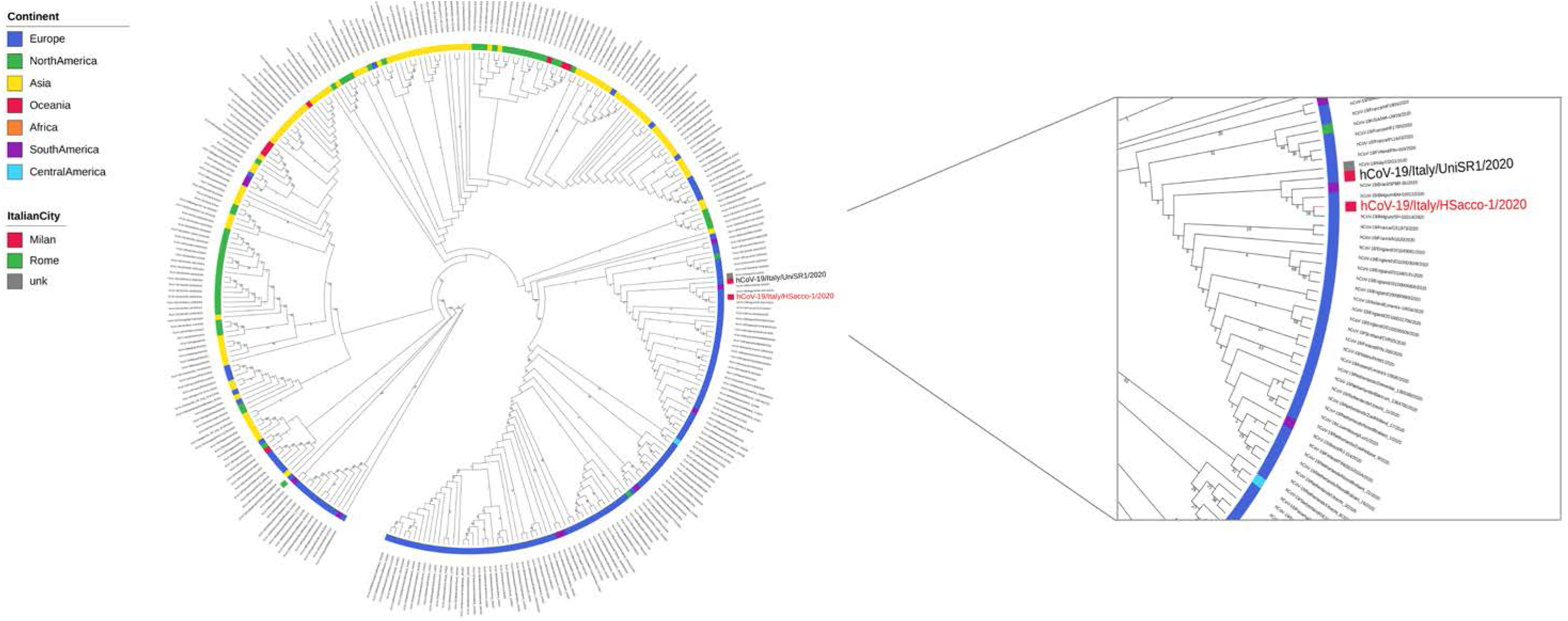
Maximum Likelihood phylogenetic tree including the the Hsacco-1 strain sequenced in this work and the 300 most similar SARS-CoV-2 strains retrieved from GISAID database. Geographic strains metadata, as retrieved from GISAID database, were mapped on the tree: the isolation continent is reported on the inner circle and, for the Italian strains only, the isolation city is reported on the external circle. The labels of the strains isolated in Milan are reported with larger size and the strain sequenced in this work is coloured in red.

In conclusion, this study firstly detected the presence of SARS CoV2 RNA in WWs and receiving rivers in the Milano Metropolitan Area, by using rapid protocols developed for diagnostic survey. Wastewater-based epidemiology for SARS CoV2 appears as a promising tool for epidemic trend monitoring to complement current clinical data. The presence of SARS-CoV-2 genome in rivers indicated the partial efficiency of the current sewerage system of the Milano Metropolitan Area, presenting similar features to Europe and USA. However, test for vitality indicated that pathogenicity of virus in wastewaters and surficial waters is negligible, and risks for public health should not be significant.

## Data Availability

All data referred to this manuscript are available upon request to the authors

## 4. Acknowledgments

Sara Giordana Rimoldi and Fabrizio Stefani equally contributed to this paper. The authors want to thanks the managers of the WWTPs for having contributed to this research.

